# HOME HEALTHCARE NEEDS AMONG OLDER ADULTS: DEMOGRAPHIC, CLINICAL, AND PSYCHOSOCIAL FACTORS

**DOI:** 10.1101/2025.07.18.25329170

**Authors:** Tran Dinh Trung, Ngo Thi Bich Ngoc, Hoang Kim Thien, Tran Thi Diep Ha, Tran Anh Quoc, Hoang Nguyen Nhat Linh, Hoang Huu Hai, Hoang Thach Thao, Nguyen Thi Mien Ha, Truong Le Khanh Dan, Hoang Thanh Huong, Trinh Ngoc Tan, Nguyen Khac Minh

**Affiliations:** Faculty of Public Health, Da Nang University of Medical and Pharmacy, Danang city, Vietnam; Rollins School of Public Health, Emory University, Geogia, USA

**Author notes:** **Corresponding author** Tran Dinh Trung Faculty of Public Health, Da Nang University of Medical Technology and Pharmacy, Danang City, Viet Nam, 500000.

**Keywords:** home healthcare, older adults, cardiovascular disease, adaptability, Vietnam

## Abstract

**Background:** Vietnam is experiencing rapid population aging, creating increased demand for healthcare services tailored to older adults. This study aimed to determine the need for home healthcare services among older adults in Da Nang and identify key demographic, clinical, and psychosocial factors associated with this need.

**Methods:** A cross-sectional study was conducted from January to May 2023 involving 667 adults aged ≥60 years in Da Nang City, Vietnam. Multi-stage sampling was used to select participants from three districts. Data were collected through face-to-face interviews using a structured questionnaire covering demographic characteristics, health status, chronic diseases, and psychosocial factors. Multivariate logistic regression was used to identify independent predictors of home healthcare service needs.

**Results:** The majority of participants (82.9%) expressed a need for home healthcare services. Significant independent predictors included advanced age (OR=2.88, 95% CI:1.51-5.49 for >70 years), female gender (OR=1.94, 95% CI:1.19-3.15), education below high school level (OR=2.60, 95% CI:1.54-4.40), cardiovascular disease (OR=2.89, 95% CI:1.59-5.24), and adaptability to new healthcare services (OR=2.95, 95% CI:1.80-4.85). The presence of two chronic diseases significantly increased the likelihood of needing home healthcare compared to having none (OR=4.06, 95% CI:1.55-10.61).

**Conclusions:** There is substantial need for home healthcare services among older adults in Da Nang, influenced by a complex interplay of demographic, health-related, and psychosocial factors. Adaptability to new healthcare services emerged as the strongest predictor alongside age, gender, education, and cardiovascular disease status. These findings can inform evidence-based policy and program development to better meet the healthcare needs of Vietnam’s aging population while supporting their preferences to age in place.

## 1. Introduction

Rapid population aging in Vietnam presents significant challenges to healthcare systems, particularly regarding the provision of appropriate care for older adults. With the proportion of people aged 60 and above expected to increase from 10.6% in 2019 to 15.5% by 2040 (1), Vietnam faces an urgent need to develop sustainable healthcare models that address the complex needs of its aging population (2, 3). Home healthcare services represent a promising approach to meeting these needs while supporting older adults’ preferences to age in place, yet research on the demand for such services and their determinants remains limited in the Vietnamese context (4).

The Vietnamese healthcare system, characterized by a mix of public and private providers, has historically emphasized facility-based care with limited attention to home-based services (5). While the Ministry of Health has recognized the importance of developing home healthcare in its recent strategic plans, implementation remains inconsistent, with significant gaps in understanding the specific needs of older adults and the factors that influence their preferences for home-based services (6). International evidence suggests that multiple factors influence home healthcare needs and utilization patterns among older adults, including demographic characteristics, health status, and psychosocial factors (7). While studies from other countries have identified advanced age, female gender, chronic disease burden, and functional limitations as predictors of home healthcare needs (8, 9), the applicability of these findings to the Vietnamese context requires careful examination due to distinct cultural, social, and healthcare system characteristics (10).

This study aims to address this knowledge gap by investigating the demand for home healthcare services among older adults in Vietnam and identifying key factors associated with this demand. Specifically, we examine the influence of demographic characteristics, health status including chronic disease burden, and psychosocial factors on the expressed need for home healthcare services. Understanding these relationships is crucial for developing evidence-based policies and programs that effectively address the healthcare needs of Vietnam’s rapidly aging population while optimizing resource allocation within the healthcare system (11). By providing insights into the complex determinants of home healthcare needs among older adults in an urban Vietnamese setting, this study contributes to the growing body of literature on aging and healthcare in Southeast Asia while offering practical implications for service development and implementation. The findings from this research can inform the design of responsive, contextually appropriate home healthcare models that align with the preferences and needs of older adults in Vietnam’s evolving healthcare landscape (12, 13).

## 2. Materials and Methods

### 2.1 Study design

This cross-sectional study was conducted from January to May 2023 in Da Nang City, Vietnam, following approval from the Ethics Committee of the Da Nang University of Medical Technology and Pharmacy, with all participants providing informed consent prior to enrollment.

### 2.2 Sampling and study population

A multi-stage sampling method was employed, first purposively selecting three districts/communes (Hai Chau, Son Tra, and Hoa Vang) from Da Nang City, then randomly selecting two wards/villages from each (Hai Chau II, Hoa Thuan Dong, Tho Quang, An Hai Bac, Hoa Tien, and Hoa Nhon), and finally conveniently selecting participants from each ward/village health center’s list of older adults until reaching the required sample size, which was calculated using the formula (1), resulting in a minimum of 606 participants, with 667 ultimately recruited after accounting for potential dropouts. Inclusion criteria were age ≥ 60 years, residence in Da Nang City for ≥1 year, and willingness to participate, while exclusion criteria included inability to hear or speak, diagnosed mental disorders, and acute life-threatening conditions.

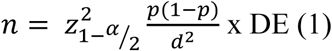

### 2.3 Data collection

Face-to-face interviews were conducted by trained population officers from local health stations, with each interview lasting approximately 10-15 minutes and taking place either at the participant’s home or the local health station, using a questionnaire that had been pilot-tested on 30 participants at Thach Thang Ward Health Station to assess feasibility and reliability before making necessary modifications.

### 2.4 Measurement

The structured questionnaire gathered demographic information (age, gender, ethnicity, occupation, education, marital status, income, economic status, and health insurance), health status details (chronic diseases, medication use, health concerns, and functional status across five domains: mobility, chewing, hearing, speaking, and vision), needs assessment for home healthcare services (including specific service types like regular health check-ups, home doctor visits, telehealth connections, nursing care, and emergency transportation), and psychosocial factors (adaptability to new healthcare services, treatment adherence, desire for guidance, awareness, support from healthcare workers, information-seeking behavior, health anxiety, skepticism about effectiveness, prior service utilization, social media usage, and service cost perception), with social support measured using the validated 12-item Multidimensional Scale of Perceived Social Support (MSPSS) that demonstrated good internal consistency (Cronbach’s alpha = 0.85).

### 2.5 Statistical analysis

Data were entered and analyzed using R-studio, with descriptive statistics presented as frequencies and percentages for categorical variables and means and standard deviations for continuous variables, while associations between the need for home healthcare services and independent variables were examined using chi-square or Fisher’s exact tests as appropriate. Three progressive multivariate logistic regression models were built (Model 1 with demographic factors, Model 2 adding health-related factors, and Model 3 incorporating psychosocial factors) to identify independent predictors of home healthcare service needs, with odds ratios and 95% confidence intervals calculated and model fit assessed using McFadden’s Pseudo R² and Akaike Information Criterion (AIC), setting statistical significance at *p < 0.05*.

## 3. Results

### 3.1 Demographic characteristics and health status

As shown in Table 1, our study included 667 older adults in Da Nang city, with a slightly higher proportion of female participants (52.3%) than males (47.7%). The largest age group was 65-70 years (41.5%), followed by those over 70 years (30.9%) and under 65 years (27.6%). Educational attainment varied considerably, with 38.4% having completed vocational training, college, or university education, while 33.0% had secondary school education, 15.6% had high school education, and 13.0% had primary school education or lower. The health profile of participants revealed that a substantial majority (79.3%) suffered from at least one chronic disease, as detailed in Table 1. Among all participants, 56.4% had one chronic condition, 11.7% had two conditions, and 11.1% had three or more conditions. Cardiovascular diseases were most prevalent (39.5%), followed by musculoskeletal disorders (28.4%), gastrointestinal diseases (21.9%), diabetes mellitus (16.8%), and respiratory conditions (16.3%). In terms of functional capacity, more than one-third of participants (36.1%) reported experiencing difficulties in daily activities. When asked about health satisfaction, only 32.7% expressed satisfaction with their health status, while 51.9% felt neutral and 15.4% were dissatisfied. Similarly, 38.0% rated their quality of life as good, 52.3% as moderate, and 9.7% as poor.

**Table 1:** Demographic characteristics and health status of study participants (n=667)

| **Characteristics** | **n** | **%** |
| --- | --- | --- |
| **Age group** |  |  |
| < 65 years | 184 | 27.6 |
| 65-70 years | 277 | 41.5 |
| > 70 years | 206 | 30.9 |
| **Gender** |  |  |
| Male | 318 | 47.7 |
| Female | 349 | 52.3 |
| **Educational level** |  |  |
| Primary school or lower | 87 | 13.0 |
| Secondary school | 220 | 33.0 |
| High school | 104 | 15.6 |
| Vocational/College/University/Postgraduate | 256 | 38.4 |
| **Chronic diseases** |  |  |
| Have chronic disease(s) | 529 | 79.3 |
| No chronic disease | 138 | 20.7 |
| **Number of chronic diseases** |  |  |
| 0 disease | 138 | 20.7 |
| 1 disease | 376 | 56.4 |
| 2 diseases | 78 | 11.7 |
| 3 diseases | 51 | 7.6 |
| 4 diseases | 21 | 3.1 |
| 5 diseases | 3 | 0.4 |
| **Types of chronic diseases\*** |  |  |
| Cardiovascular disease | 209 | 39.5 |
| Musculoskeletal disease | 150 | 28.4 |
| Gastrointestinal disease | 116 | 21.9 |
| Diabetes mellitus | 89 | 16.8 |
| Respiratory disease | 86 | 16.3 |
| Hepatobiliary and pancreatic disease | 40 | 7.6 |
| Other diseases | 39 | 7.4 |
| Neurological disease | 36 | 6.8 |
| Renal disease | 19 | 3.6 |
| **Functional status** |  |  |
| With difficulties | 241 | 36.1 |
| Without difficulties | 426 | 63.9 |
| **Health satisfaction level** |  |  |
| Not satisfied | 103 | 15.4 |
| Neutral | 346 | 51.9 |
| Satisfied | 218 | 32.7 |
| **Quality of life assessment** |  |  |
| Poor | 65 | 9.7 |
| Moderate | 349 | 52.3 |

| Characteristics | n | % |
| --- | --- | --- |
| Good | 253 | 38.0 |

\*Percentages calculated based on total number of participants with chronic diseases (n=529)

### 3.2 Psychosocial characteristics

Table 2 illustrates the psychosocial profile of the participants. A majority (65.7%) considered themselves adaptable to new healthcare services, and 86.5% reported adhering to treatment recommendations. Most participants (78.3%) expressed a desire for guidance in navigating home healthcare services. Awareness and utilization of home healthcare services showed mixed results. While 56.7% of participants were aware of such services, only 47.7% had actively sought information about them, and just 40.8% had previously utilized these services. Despite 62.5% reporting support from healthcare workers, 36.0% remained skeptical about the effectiveness of home healthcare services. Health anxiety was common among participants, with 79.8% reporting concerns about their health. Regarding social connections, 68.1% reported moderate social support and 31.9% reported high support levels. More than half (53.7%) used social media platforms. Notably, 67.2% perceived home healthcare services as more expensive than hospital visits.

**Table 2:** Psychosocial characteristics of study participants (n=667)

| Psychosocial factors | Classification | n (%) |
| --- | --- | --- |
| **Adaptability** |  |  |
|  | Adaptable | 438 (65.7) |
|  | Not adaptable | 229 (34.3) |
| **Treatment adherence** |  |  |
|  | Adherent | 577 (86.5) |
|  | Non-adherent | 90 (13.5) |
| **Desire for guidance** |  |  |
|  | Desires guidance | 522 (78.3) |
|  | Does not desire guidance | 145 (21.7) |
| **Awareness of home healthcare services** |  |  |
|  | Aware | 378 (56.7) |
|  | Unaware | 289 (43.3) |
| **Support from healthcare workers** |  |  |
|  | With support | 417 (62.5) |
|  | Without support | 250 (37.5) |
| **Service information seeking** |  |  |
|  | Has sought information | 318 (47.7) |

| **Psychosocial factors** | **Classification** | **n (%)** |
| --- | --- | --- |
|  | Has not sought information | 349 (52.3) |
| **Health anxiety** |  |  |
|  | Anxious | 532 (79.8) |
|  | Not anxious | 135 (20.2) |
| **Skepticism about effectiveness** |  |  |
|  | Skeptical | 240 (36.0) |
|  | Not skeptical | 427 (64.0) |
| **Prior service utilization** |  |  |
|  | Never used | 395 (59.2) |
|  | Has used | 272 (40.8) |
| **Social support** |  |  |
|  | Moderate support | 454 (68.1) |
|  | High support | 213 (31.9) |
| **Social media usage** |  |  |
|  | Uses social media | 358 (53.7) |
|  | Does not use social media | 309 (46.3) |
| **Service cost perception** |  |  |
|  | Higher than hospital visit | 448 (67.2) |
|  | Lower than hospital visit | 219 (32.8) |

### 3.3 Chronic disases associated with the need for home healthcare services

Table 3 presents the relationship between chronic diseases and the need for home healthcare services. After adjusting for age, gender, and educational level, having cardiovascular disease significantly increased the likelihood of needing home healthcare services (OR=2.74, 95% CI: 1.55-4.87, *p<0.05*). Interestingly, those with gastrointestinal diseases showed lower odds of needing these services (OR=0.45, 95% CI: 0.27-0.73, *p<0.05*), while those with hepatobiliary and pancreatic diseases had significantly higher odds (OR=4.41, 95% CI: 1.01-19.16, *p<0.05*). Having two chronic diseases was significantly associated with higher need compared to having none (OR=4.06, 95% CI: 1.55-10.61, *p<0.05*).

**Table 3:**
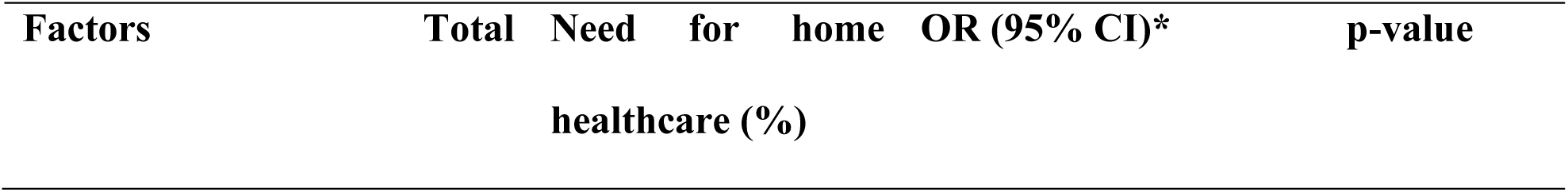
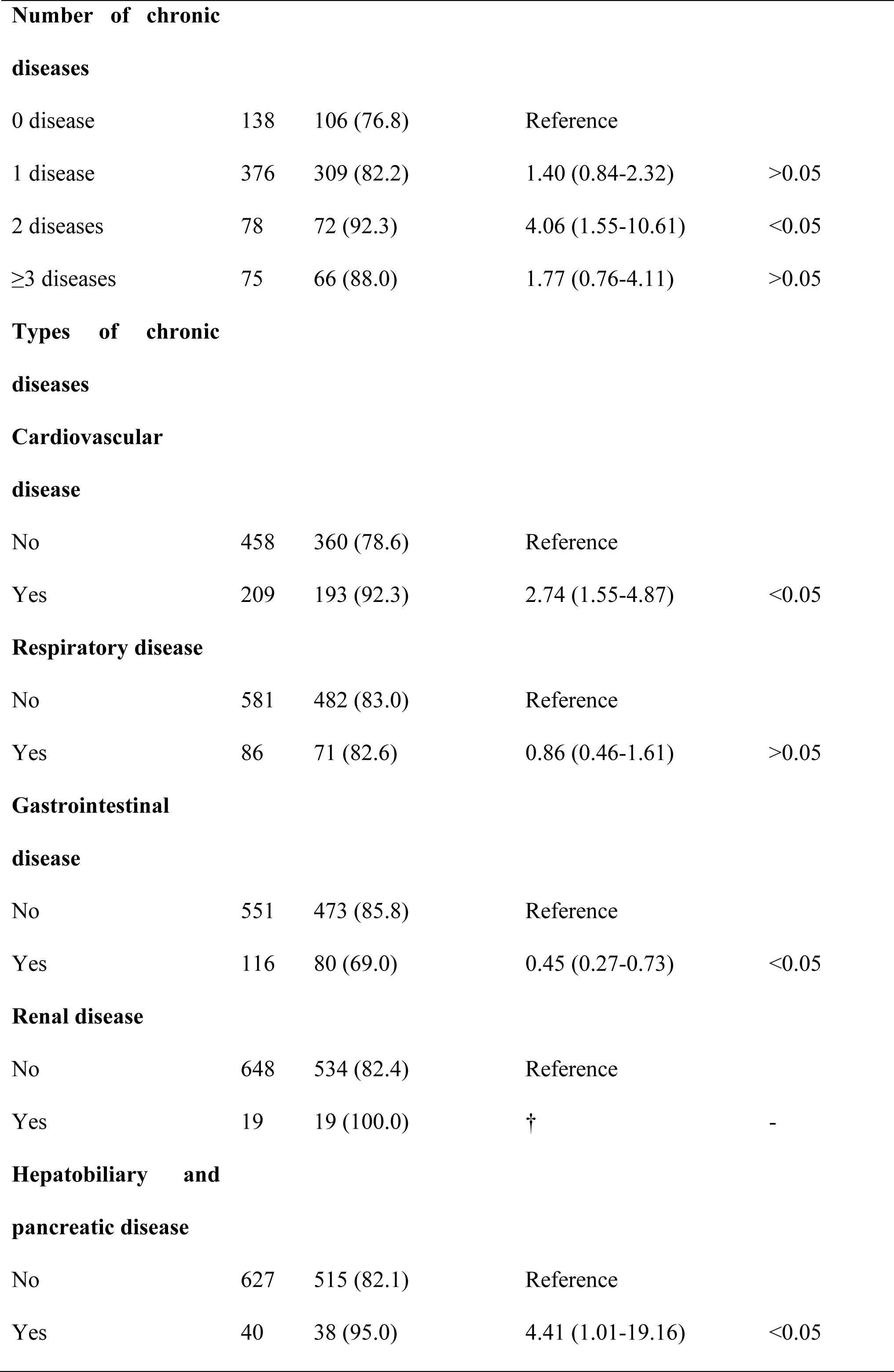
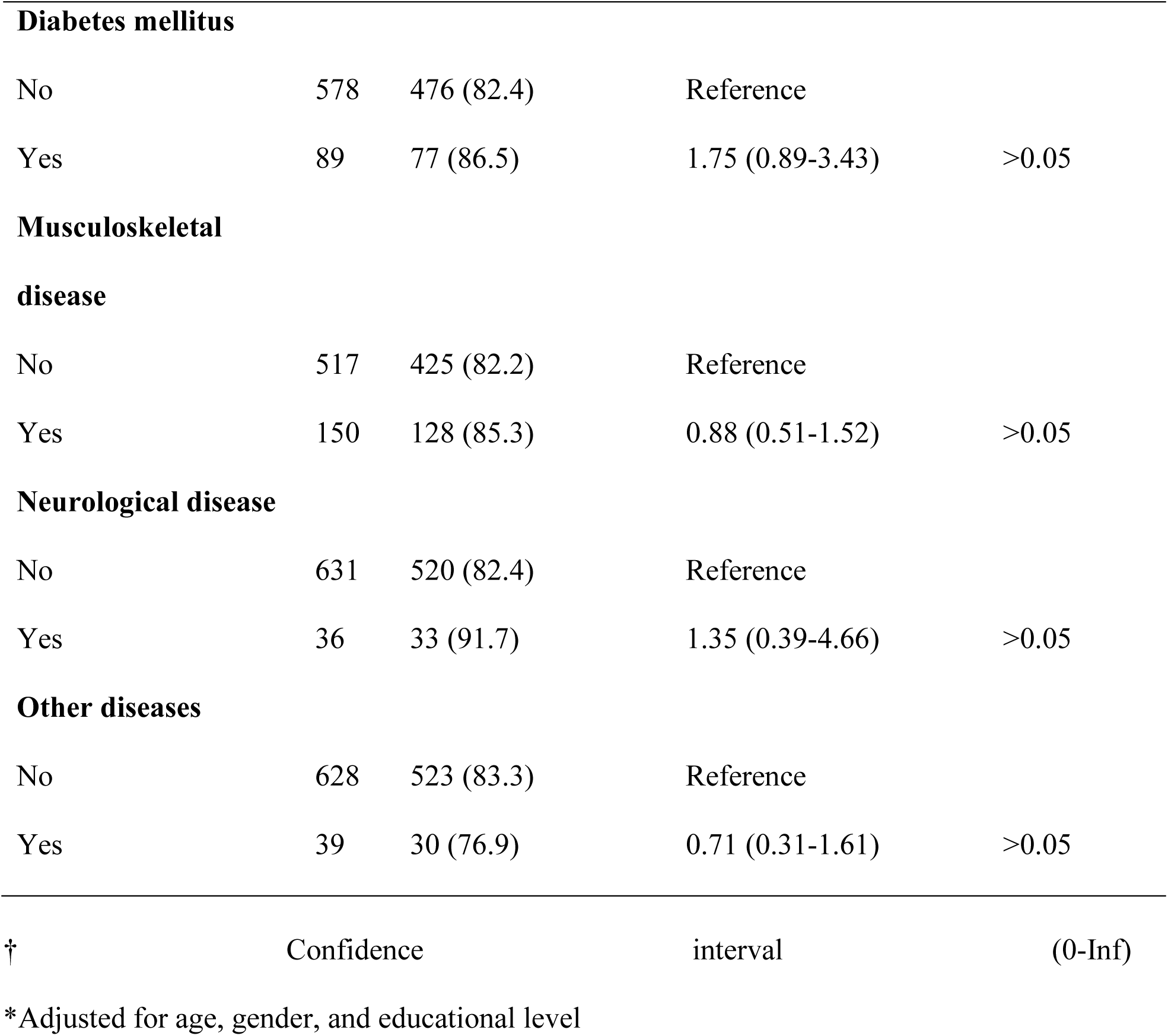
Association between chronic diseases and the need for home healthcare services.

### 3.3 Psychosocial factors associated with the need for home healthcare services

As shown in Table 4, several psychosocial factors were significantly associated with the need for home healthcare services. Participants who were adaptable to new healthcare services had higher odds of expressing need (OR=3.69, 95% CI: 2.35-5.78, *p<0.05*) compared to those who were not adaptable. Similarly, those receiving support from healthcare workers showed greater need (OR=2.11, 95% CI: 1.37-3.25, p<0.05), as did those using social media (OR=2.33, 95% CI: 1.46-3.72, *p<0.05*).

**Table 4:** Association between psychosocial factors and the need for home healthcare services.

| **Psychosocial factors** | **Total** | **Need for home healthcare (%)** | **OR (95% CI)\*** | **P-value** |
| --- | --- | --- | --- | --- |
| **Adaptability** |  |  |  |  |
| Not adaptable | 229 | 164 (71.6) | Reference |  |
| Adaptable | 438 | 389 (88.8) | 3.69 (2.35-5.78) | **<0.05** |
| **Support from healthcare workers** |  |  |  |  |
| Without support | 250 | 191 (76.4) | Reference |  |
| With support | 417 | 362 (86.8) | 2.11 (1.37-3.25) | **<0.05** |
| **Health anxiety** |  |  |  |  |
| Not anxious | 135 | 96 (71.1) | Reference |  |
| Anxious | 532 | 457 (85.9) | 1.52 (0.93-2.47) | >0.05 |
| **Social media usage** |  |  |  |  |
| Does not use | 309 | 252 (81.6) | Reference |  |
| Uses social media | 358 | 301 (84.1) | 2.33 (1.46-3.72) | **<0.05** |
| **Social support** |  |  |  |  |
| Moderate support | 344 | 284 (82.6) | Reference |  |
| High support | 323 | 269 (83.3) | 1.05 (0.66-1.69) | >0.05 |

\*Adjusted for age, gender, and educational level
P-value represents statistical significance of adjusted OR

### 3.4 Predicting models for the need for home healthcare services

Table 5 presents three progressive multivariate regression models predicting the need for home healthcare services. The fully adjusted Model 3, which incorporated demographic, health, and psychosocial factors, demonstrated the best fit (McFadden’s Pseudo R²=0.191, AIC=515.62). In this comprehensive model, several factors emerged as significant independent predictors of the need for home healthcare services: age over 70 years (OR=2.88, 95% CI: 1.51 – 5.49), age 65-70 years (OR=2.17, 95% CI: 1.29 – 3.64), female gender (OR=1.94, 95% CI: 1.19 – 3.15), education below high school level (OR=2.60, 95% CI: 1.54 – 4.40), cardiovascular disease (OR=2.89, 95% CI: 1.59-5.24), and adaptability to new healthcare services (OR=2.95, 95% CI: 1.80 – 4.85). These findings highlight that the need for home healthcare services among older adults in Da Nang is influenced by a complex interplay of demographic characteristics, health conditions, and psychosocial factors, with adaptability to new healthcare services emerging as the strongest predictor alongside age, gender, education, and cardiovascular disease status.

**Table 5:** Multivariate regression model predicting the need for home healthcare services in Da Nang

| **Factors** | **Model 1**<br>**(Demographic factors)** | **Model 2 (+ Health factors)** | **Model 3 (+ Psychosocial factors)** |
| --- | --- | --- | --- |
| **Age group** |  |  |  |
| < 65 years | Reference | Reference | Reference |
| 65-70 years | 2.33 (1.45 – 3.75) | 2.28 (1.4 – 3.72) | 2.17 (1.29 – 3.64) |
| > 70 years | 2.95 (1.63 – 5.35) | 2.84 (1.54 – 5.26) | 2.88 (1.51 – 5.49) |
| **Female** | 2.16 (1.37 – 3.4) | 2.24 (1.41 – 3.56) | 1.94 (1.19 – 3.15) |
| **Below High school** | 2.32 (1.47 – 3.64) | 2.07 (1.29 – 3.31) | 2.6 (1.54 – 4.4) |
| **Cardiovascular disease** |  | 2.87 (1.61 – 5.12) | 2.89 (1.59 – 5.24) |
| **Hepatobiliary and pancreatic disease** |  | 4.94 (1.1 – 22.16) | 2.92 (0.64 – 13.35) |
| **Diabetes mellitus** |  | 1.42 (0.71 – 2.82) | 1.2 (0.58 – 2.47) |
| **Adaptability** |  |  | 2.95 (1.8 – 4.85) |
| **Support from healthcare workers** |  |  | 1.2 (0.72 – 2.01) |
| **Social media usage** |  |  | 1.51 (0.91 – 2.52) |
| **McFadden's Pseudo R<sup>2</sup>** | 0.102 | 0.140 | 0.191 |
| **AIC** | 557.34 | 540.92 | 515.62 |

## 4. Discussion

This study aimed to determine the need for home healthcare services among older adults and identify key factors associated with this need. Our findings revealed that 82.9% of older adults expressed a need for home healthcare services, with independent predictors including advanced age, female gender, lower education level, cardiovascular disease, and adaptability to new healthcare services. The significance and novelty of the study lie in (1) its comprehensive examination of the complex interplay between demographic, commorbidities, and psychosocial factors influencing home healthcare needs in Vietnam’s rapidly aging urban population, and (2) its identification of adaptability to new healthcare services as the strongest psychosocial predictor, offering actionable insights for developing targeted healthcare policies and service delivery models that can better meet the needs of older adults in this emerging healthcare context.

First, our findings demonstrate that demographic factors significantly predict the need for home healthcare services among older adults, with advanced age, female gender, and lower education emerging as strong predictors. These results align with international studies reporting similar demographic patterns in healthcare utilization. In rural Vietnam, advanced age, higher education, and female gender have been associated with increased prevalence of chronic conditions requiring ongoing care (14). This observation is consistent with global trends where aging populations experience higher rates of multimorbidity necessitating home-based services. The gender disparity observed in our study mirrors findings from studies of frailty and healthcare utilization where women consistently demonstrate higher healthcare needs, potentially due to longer life expectancy and higher prevalence of disabling conditions (15, 16). The association between lower education and increased home healthcare needs reflects broader social determinants of health, where educational status influences health literacy, preventive health behaviors, and timely access to healthcare services (17). These associations can be explained through multiple mechanisms, including the cumulative disadvantage theory where socioeconomic inequalities compound throughout the life course, leading to greater healthcare dependencies in later life (15, 16). From a practical perspective, these findings suggest that home healthcare services in Vietnam’s rapidly aging population should incorporate targeted approaches for the oldest-old, particularly women and those with limited formal education, with emphasis on culturally appropriate health education and support services that address the specific barriers these demographic groups face (13).

The identification of CVD as the strongest health-related predictor of home healthcare needs underscores the critical role of chronic disease burden in shaping care requirements among older adults (14–16, 18–20). This finding aligns with global epidemiological patterns where CVD accounts for nearly one-third of all disability-adjusted life years in individuals over 70, necessitating complex care coordination (18, 21). Comparative analysis reveals that while diabetes and respiratory conditions showed weaker associations in our cohort, the prominence of CVD mirrors outcomes observed in South African managed-care populations, where comorbid CVD and depression doubled hospitalization rates for macrovascular complications compared to non-depressed counterparts (19). The pathophysiological basis for this association lies in CVD’s dual impact on functional capacity and care complexity—progressive cardiac dysfunction limits mobility, while polypharmacy requirements (averaging 11.6 medications in U.S. cohorts) elevate risks of drug interactions and adherence challenges that home healthcare teams are uniquely positioned to address (21, 22). Mechanistically, the interplay between CVD-related physiological decline and psychosocial factors creates a self-reinforcing cycle of dependency (23, 24). Impaired cardiac output and exercise intolerance, demonstrated by 2-minute step test performance declines in home care patients, reduce capacity for self-care activities while increasing fall risks (25–27). Concurrently, CVD management protocols requiring strict blood pressure monitoring and anticoagulant titration demand clinical oversight that traditional outpatient models struggle to provide, as evidenced by 18.% to 23.6% 30-day readmission rates in U.S. homebound CVD patients (28). High polypharmacy prevalence and severe drug-drug interaction potential among older cardiovascular disease patients necessitate effective home healthcare strategies (21). Medication review services and patient education can identify inappropriate prescriptions and medication problems (29). Vigilant drug record review and therapy adjustment are crucial to prevent adverse reactions (21). Furthermore, integrating guided physical activity programs tailored to cardiac limitations could break the inertia-adherence cycle, leveraging evidence that supervised home-based exercise improves functional status in CVD patients (18).

The identification of adaptability to new healthcare services as the strongest psychosocial predictor reveals a critical dimension in care utilization patterns among older adults. This finding aligns with emerging research on technology adoption in aging populations, where digital adaptability and openness to novel care modalities significantly predict engagement with telehealth platforms and remote monitoring systems (30, 31). Comparative analysis with international cohorts demonstrates that psychological flexibility—the capacity to adjust behaviors to align with healthcare protocols—mediates care adherence more powerfully than static personality traits like resilience or optimism (32, 33). For example, previous study of digital leisure adoption found older adults with high adaptability scores exhibited 84.4% higher service utilization rates despite lower baseline technological literacy (34), mirroring the behavioral patterns observed in our cohort. Mechanistically, adaptability operates through dual pathways: cognitive reframing of healthcare interactions (reducing perceived intrusiveness of home visits) and enhanced problem-solving during care transitions (mitigating disruptions to daily routines) (24, 33). Neurobiological correlates further elucidate this relationship, as functional MRI studies associate adaptability with increased prefrontal cortex activation during decision-making tasks—a neural signature linked to improved integration of complex care instructions (35). Conversely, rigid cognitive patterns observed in low-adaptability individuals correlate with amygdala hyperactivity, potentiating care-related anxiety and avoidance behaviors (32, 36). These neurocognitive insights intersect with sociological frameworks positing that adaptability buffers against the “technological alienation” often experienced by older adults navigating digitized care systems (37, 38). Concurrently, service protocols should incorporate adaptability assessments during intake evaluations, enabling tailored care plans that progressively introduce novel interventions while respecting established routines (39, 40). This dual-strategy approach addresses both individual capacity building and systemic barriers, creating pathways for sustainable psychosocial integration within Vietnam’s evolving home healthcare infrastructure (41–43).

Several limitations should be acknowledged in this study. First, its cross-sectional design prevents establishing causal relationships between identified factors and home healthcare needs. Second, convenience sampling from health station lists may have introduced selection bias by potentially overrepresenting older adults already engaged with healthcare services, while underrepresenting socially isolated individuals with potentially greater needs. Third, self-reported health measures and retrospective information on service utilization may be subject to recall bias. Additionally, despite the comprehensive approach to measuring psychosocial factors, more nuanced cultural dimensions specific to Vietnamese aging experiences, such as filial piety expectations and traditional healing preferences, warrant deeper exploration. Future research should employ longitudinal designs to track how needs evolve over time, include cost-effectiveness analyses of various home healthcare models within Vietnam’s mixed public-private healthcare system, and investigate implementation barriers through qualitative approaches with stakeholders including healthcare providers, policymakers, and family caregivers.

## 5. Conclusion

This study provides an evidence of substantial need for home healthcare services among older adults. The research identifies a constellation of predictors including advanced age, female gender, lower education, cardiovascular disease, and notably, adaptability to new healthcare services—offering valuable insights for developing responsive and contextually appropriate home healthcare systems. These findings have significant implications for healthcare policy and practice in Vietnam’s rapidly aging society, suggesting that successful implementation of home healthcare services requires not only addressing clinical needs but also engaging with psychosocial factors that influence service acceptability and utilization. As Vietnam navigates population aging alongside socioeconomic transformation, these insights can inform evidence-based policy and program development that aligns with older adults’ preferences to age in place while ensuring high-quality, accessible healthcare that meets their complex and evolving needs.

## Data Availability

All data produced in the present work are contained in the manuscript

## Acknowledgements

The authors express sincere gratitude to all older adults who participated in this study, as their valuable time and willingness to share their experiences made this research possible. Special appreciation goes to Dr. Tran Minh Huan for providing critical feedback and insightful comments during manuscript revision. The proofreading process was assisted by ChatGPT, and the authors take full responsibility for the content after proofreading.

## Authors’ contributions

**TDT and NKM** contributed to the completion of interpretation of the data and the manuscript. **NTBN, HKT, TDDH, HNNL, TAQ, HTH, TLKD, TNT** contributed to clinical data collection. **TDT, NTMH** contributed to critical revision of the manuscript for important intellectual content. All authors have read and approved the final manuscript.

